# COVID-19 mRNA vaccines induce robust levels of IgG but limited amounts of IgA within the oronasopharynx of young children

**DOI:** 10.1101/2024.04.15.24305767

**Authors:** Ying Tang, Brittany P. Boribong, Zoe N. Swank, Melina Demokritou, Maria A.F. Luban, Alessio Fasano, Michelle Du, Rebecca L. Wolf, Joseph Griffiths, John Shultz, Ella Borberg, Sujata Chalise, Wanda I. Gonzalez, David R. Walt, Lael M. Yonker, Bruce H. Horwitz

**Affiliations:** Division of Gastroenterology, Hepatology, and Nutrition, Boston Children’s Hospital, Boston, MA 02115, USA; Harvard Medical School, Boston, MA 02115, USA; Mucosal Immunology and Biology Research Center, Massachusetts General Hospital, Boston, MA 02114, USA; Department of Pediatrics, Massachusetts General Hospital, Boston, MA 02114, USA; Department of Pathology, Brigham and Women’s Hospital, Boston, MA 02115, USA; Wyss Institute for Biologically Inspired Engineering, Harvard University, Boston, MA 02215, USA; Division of Emergency Medicine, Boston Children’s Hospital, Boston, MA 02115, USA

## Abstract

**Key points:**

- Current COVID-19 mRNA vaccine induces salivary and nasal SARS-CoV-2 specific IgG but not IgA production in children under 5 years of age
- Mucosal anti-spike IgA is important for immune complex-mediated neutrophil extracellular trap formation against SARS-CoV-2 in the airway

**Background:** Understanding antibody responses to SARS-CoV-2 vaccination is crucial for refining COVID-19 immunization strategies. Generation of mucosal immune responses, including mucosal IgA, could be of potential benefit to vaccine efficacy, yet limited evidence exists regarding the production of mucosal antibodies following the administration of current mRNA vaccines to young children.

**Methods:** We measured the levels of antibodies against SARS-CoV-2 from a cohort of children under 5 years of age undergoing SARS-CoV-2 mRNA vaccination (serially collected, matched serum and saliva samples, N=116) or on convenience samples of children under 5 years of age presenting to a pediatric emergency department (nasal swabs, N=103). Further, we assessed salivary and nasal samples for the ability to induce SARS-CoV-2 spike-mediated neutrophil extracellular traps (NET) formation.

**Results:** Longitudinal analysis of post-vaccine responses in saliva revealed the induction of SARS-CoV-2 specific IgG but not IgA. Similarly, SARS-CoV-2 specific IgA was only observed in nasal samples obtained from previously infected children with or without vaccination, but not in vaccinated children without a history of infection. In addition, oronasopharyngeal samples obtained from children with prior infection were able to trigger enhanced spike-mediated NET formation, and IgA played a key role in driving this process.

**Conclusions:** Despite the induction of specific IgG in the oronasal mucosa, current intramuscular vaccines have limited ability to generate mucosal IgA in young children. These results confirm the independence of mucosal IgA responses from systemic humoral responses following mRNA vaccination and suggest potential future vaccination strategies for enhancing mucosal protection in this young age group.

## INTRODUCTION

While there is clear evidence that current COVID-19 mRNA vaccines induce robust and protective systemic immune responses, the ability of these vaccines to induce mucosal responses is less understood. Mucosal immune responses may provide additive benefits potentially important for limiting transmission and increasing effectiveness against severe disease [1]. It has been demonstrated in animal models that targeted nasal immunization, but not intramuscular immunization, with ChAd-SARS-CoV-2 induces robust mucosal anti-IgA responses with near sterilizing immunity, suggesting a role for mucosal IgA responses in preventing SARS-CoV-2 infection and transmission [2]. Moreover, nasal SARS-CoV-2 specific antibody responses have been associated with lower viral loads and milder systemic symptoms of COVID-19 [3]. Studies on adults revealed that prior infection induces significantly higher mucosal IgA than mRNA vaccination [4–6], underscoring the limited impact of intramuscular vaccination on the induction of mucosal SARS-CoV-2 specific IgA in adults [7]. Young children have developing immune systems with significantly reduced capacity to generate circulating anti-SARS-CoV-2 IgA following vaccination as compared to adults [8]. However, studies examining mucosal IgA responses in children following SARS-CoV2 mRNA vaccinations are limited.

Here, we longitudinally evaluated both serological and salivary antibody responses in a cohort of children under 5 years of age with and without a prior history of SARS-CoV-2 infection following primary mRNA vaccination. We also compared antibody levels in nasal samples obtained from children with a history of COVID-19, those with a prior history of vaccination, those with both infection and vaccination, or those with neither. Additionally, we explored the ability of spike-specific mucosal antibodies to induce neutrophil activation. Our results reveal that while mRNA vaccination can generate robust systemic and mucosal IgG production, vaccination alone has a limited abillity to induce oronasopharyngeal IgA, nor does it boost mucosal IgA levels induced by prior SARS-CoV-2 infection. Further, our data also suggest that IgA produced in response to prior SARS-CoV-2 infection is a key driver of anti-SARS-CoV-2 antibody-induced neutrophilic activation.

## METHODS

### Study Design

#### Longitudinal cohort

Children aged 5 years or younger undergoing a COVID-19 mRNA vaccination series were enrolled in this study. Informed consent was obtained from parents/legal guardians. The IRB of Massachusetts General Hospital gave ethical approval for this work. SARS-CoV-2 infection history and demographic information were obtained from electronic medical records. Samples from individuals who were infected during the vaccine series were excluded from this analysis. All subjects received either Pfizer (BNT162b2) or Moderna (mRNA-173) for primary vaccine doses. Samples were collected before vaccination (V0) and 2-4 weeks following the first, the second, and (in those receiving the Pfizer vaccine) the third vaccine doses (V1, V2, V3, respectively). Saliva was collected by holding a SalivaBio swab (Salimetrics) under the tongue for 2 minutes or until fully saturated. The saturated swab was then placed in the upper chamber of the Swab Storage Tube (Salimetrics) and centrifuged at 450g at 4°C for 15 minutes. Saliva was collected, aliquoted, and stored at −80°C until use. Blood was collected via venipuncture into serum separation tubes (BD) or by a microneedle capillary blood collection device. Serum was collected, aliquoted, and stored at −80°C until use.

#### Emergency department convenience cohort

Children under 5 years old presenting to the Emergency Department at Boston Children’s Hospital (BCH) were enrolled in this study. Written informed consent was acquired from parents/legal guardians. The IRB of Boston Children’s Hospital gave ethical approval for this work. Participants with a current positive SARS-CoV-2 PCR test were excluded from this study, and their vaccination status, prior infection status as well as demographic information were obtained from a parental questionnaire. Following completion of clinically indicated viral testing employing a nasopharyngeal swab, discarded viral transport medium (VTM) was retrieved and stored at −80°C until use.

### Simoa anti-SARS-CoV-2 antibody measurements

Saliva samples were diluted 64-fold in StartingBlockTM T20 blocking buffer (Thermo Fisher Scientific) containing protease inhibitors (HaltTM Protease Inhibitor Cocktail, Thermo Fisher Scientific). Single molecule array (Simoa) assays were then used to measure anti-S1, anti-RBD, anti-spike, and anti-nucleocapsid antibodies, as previously described [9]. Briefly, using an HD-X Analyzer (Quanterix Corporation, Billerica MA), the diluted samples were incubated with dye-encoded magnetic beads coated with recombinant proteins. The beads were washed and resuspended in a solution of biotinylated anti-human-IgG antibody. The beads were then washed again and resuspended in a solution of streptavidin-conjugated β-galactosidase. Lastly, the beads were resuspended in a solution of resorufin β-D-galactopyranoside and loaded into a microwell array for imaging. Average enzymes per bead (AEB) values were calculated by the HD-X software and normalized between runs using a COVID-19 positive serum standard.

### Nasal antibody detection

VTM samples were thawed and centrifuged at 3000g for 5 mins. SARS-CoV-2 anti-S1, -S2, - RBD and -Nucleocapsid IgG and IgA levels were determined using MILLIPLEX® SARS-CoV-2 Antigen Panel 1 IgG assay (Millipore Sigma, Cat. No. HC19SERG1-85K) and MILLIPLEX® SARS-CoV-2 Antigen Panel 1 IgA assay (Millipore Sigma, Cat. No. HC19SERA1-85K), respectively. The protocol was followed as described by the manufacturer, except 50µL/well of undiluted VTM samples were used as the starting material, and an additional fixation step with 4% PFA was included following the final wash. Samples were analyzed using the Luminex^TM^ 200^TM^ system. All samples were measured in duplicate, and control beads were used for normalization.

### NETosis assay

The NETosis assay was performed as previously described [10]. Briefly, microfluidic devices were primed with RPMI media with no FBS. Neutrophils were isolated from healthy donors using the Easysep Direct Neutrophil Isolation Kit (STEMCELL Technologies). Isolated neutrophils were stained with 32 µM Hoeschst 3342 dye and mixed with SYTOX green (final concentration 2 µM). Stained neutrophils were stimulated with either pooled saliva samples or individual samples of VTM in the presence or absence of spike-coated NeutrAvidin beads. The cell suspensions were then loaded into a microfluidic device and imaged with brightfield, FITC, and DAPI fields every 10 minutes for 6 hours. NETosis was then quantified using FIJI and the TrackMate plugin.

### Statistical analysis

Two-tailed Mann-Whitney U tests were conducted to identify significant differences between groups in GraphPad Prism version 10.1. Statistical significance is defined as *p < 0.05, **p < 0.01, ***p < 0.001, and ****p < 0.0001.

## RESULTS

### mRNA vaccination fails to induce spike-specific IgA in saliva

To quantify mucosal and serologic antibody responses generated by COVID-19 mRNA vaccination, we evaluated saliva and blood samples collected from healthy children with and without prior history of COVID-19 based on their medical records (demographics shown in Table 1). Matched serum and saliva samples were collected longitudinally prior to vaccination and 4 weeks following each vaccine dose (Figure 1A). Participants were divided into two groups: “Vaccine-only” (no prior infection) and “Vaccine/Infection” (with prior SARS-CoV-2 infection).

**Figure 1.**
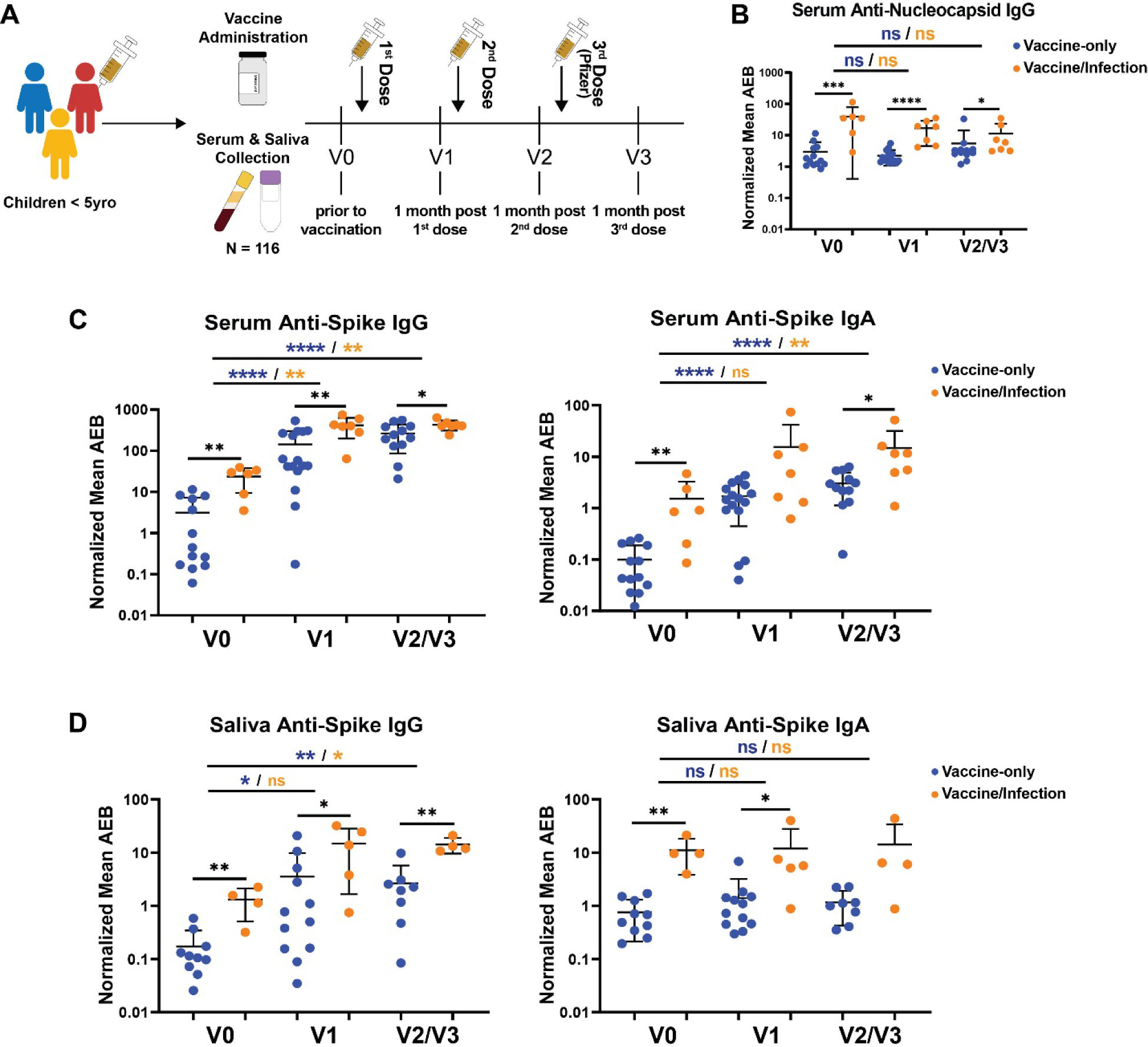
SARS-CoV-2 mRNA vaccination fails to induce spike-specific IgA in saliva. **(A)** Schematic overview of study design and sample collection timeline. **(B)** Serum anti-nucleocapsid IgG level indicates prior SARS-CoV-2 infection status. Anti-nucleocapsid IgG are shown for groups with and without a prior history of COVID-19. **(C)** Serum anti-spike IgG (left) and IgA (right) levels are shown. Differences between groups are shown as black asterisks. Differences between time points within groups are shown as blue or orange asterisks. Error bar represents the mean value and the standard deviation. Two-tailed Mann-Whitney U tests were performed between individual groups, and statistical significance is defined as *p < 0.05, **p < 0.01, ***p < 0.001, and ****p < 0.0001.

**Table 1.** Characteristics of participants in longitudinal cohort.

|  | Saliva |  |  |  | Serum |  |  |  |
| --- | --- | --- | --- | --- | --- | --- | --- | --- |
|  | V0<br>(N = 14) | V1<br>(N = 17) | V2<br>(N = 13) | V3<br>(N = 4) | V0<br>(N = 19) | V1<br>(N = 23) | V2<br>(N = 20) | V3<br>(N = 6) |
| **Age (month)** |  |  |  |  |  |  |  |  |
| Minimum | 6.87 | 6.87 | 6.87 | 6.81 | 9.24 | 6.87 | 6.87 | 6.97 |
| Median (IQR) | 12.72 (2.2) | 13.74 (15.4) | 13.05 (29) | 12.86 (10.7) | 13.74 (29.8) | 13.74 (15.6) | 14.28 (29.4) | 12.85 (26) |
| Maximum | 55.53 | 55.53 | 55.53 | 48.46 | 55.53 | 55.53 | 55.53 | 48.46 |
| **Sex** |  |  |  |  |  |  |  |  |
| Female | 7 (50%) | 8 (47.1%) | 6 (46.2%) | 2 (50%) | 8 (42.1%) | 11 (47.8%) | 10 (50%) | 4 (66.7%) |
| Male | 7 (50%) | 9 (52.9%) | 7 (53.8%) | 2 (50%) | 11 (57.9%) | 12 (52.2%) | 10 (50%) | 2 (33.3%) |
| **Race** |  |  |  |  |  |  |  |  |
| White | 11 (78.6) | 9 (52.9%) | 8 (61.5%) | 4 (100%) | 12 (63.1%) | 12 (52.2%) | 12 (60%) | 5 (83.3%) |
| Black/African American | 0 (0%) | 0 (0%) | 0 (0%) | 0 (0%) | 0 (0%) | 1 (4.3%) | 0 (0%) | 0 (0%) |
| Asian | 0 (0%) | 2 (11.8%) | 1 (7.7%) | 0 (0%) | 3 (15.8%) | 3 (13%) | 3 (15%) | 0 (0%) |
| Other/Unknown | 3 (21.4) | 6 (35.3) | 4 (30.8%) | 0 (0%) | 4 (21.1%) | 7 (30.4%) | 5 (25%) | 1 (16.7%) |
| **Ethnicity** |  |  |  |  |  |  |  |  |
| Hispanic | 0 (0%) | 0 (0%) | 0 (0%) | 4 (100%) | 0 (0%) | 1 (4.3%) | 0 (0%) | 0 (0%) |
| Not Hispanic | 9 (64.3%) | 9 (52.9%) | 8 (61.5%) | 0 (0%) | 13 (68.4%) | 13 (56.5%) | 13 (65%) | 5 (83.3%) |
| Unknown | 5 (35.7%) | 8 (47.1%) | 5 (38.5%) | 0 (0%) | 6 (31.6%) | 9 (39.1%) | 7 (35%) | 1 (16.7%) |
| **COVID-19 Vaccine Status** |  |  |  |  |  |  |  |  |
| Not vaccinated | 14 (100%) | 0 (0%) | 0 (0%) | 0 (0%) | 19 (100%) | 0 (0%) | 0 (0%) | 0 (0%) |
| Pfizer-BioNTech | 0 (0%) | 5 (29.4) | 5 (38.5%) | 4 (100%) | 0 (0%) | 15 (65.2%) | 7 (35%) | 6 (100%) |
| Moderna | 0 (0%) | 12 (70.6%) | 8 (61.5%) | 0 (0%) | 0 (0%) | 8 (34.8%) | 13 (65%) | 0 (0%) |

Consistent with the prior history, serum anti-nucleocapsid IgG levels were significantly higher in the Vaccine/Infection group than in the Vaccine-only group (Figure 1B). As expected, prior to vaccination (baseline, V0), we found significantly higher levels of anti-spike IgG and IgA in the serum of participants in the Vaccine/Infection group than in the Vaccine-only group (Figure 1C). Anti-spike IgG and IgA were significantly higher in serum collected following the completion of vaccination (V2 or V3) than prior to vaccination in both groups, although levels of both IgG and IgA remained higher in the Vaccine/Infection group than in the Vaccine-only group throughout the time course (Figure 1C). Similar patterns were observed for both anti-S1 and anti-RBD responses in serum samples (Figure S1).

Similar to responses in the serum, prior to vaccination, salivary anti-spike IgG was significantly higher in the Vaccine/Infection group than in the Vaccination-only group, and was significantly higher following completion of vaccination than prior to vaccination in both groups (Figure 1D, left). Likewise, salivary levels of anti-spike IgG remained significantly higher in the Vaccine/Infection group than in the Vaccine-only group throughout the time course, and similar patterns were observed for salivary anti-S1 and anti-RBD IgG (Figure S2A). While levels of anti-spike IgA in the saliva at baseline were also significantly higher in the Vaccine/Infection group than in the Vaccine-only group, we were unable to detect a significant increase in levels of anti-spike IgA in either group following vaccination (Figure 1D, right). Small but statistically significant increases in the levels of anti-S1 IgA but not in anti-RBD IgA were observed in the Vaccine-only group following vaccination (Figure S2B). Taken together, these observations suggest that the ability of COVID-19 mRNA vaccination to induce salivary IgA is quite limited.

### mRNA vaccination fails to induce anti-spike IgA in the nasal mucosa

To further evaluate nasopharyngeal antibody levels following mRNA vaccination and/or SARS-CoV-2 infection, we collected viral transport media (VTM) samples used for testing of material collected on nasopharyngeal swabs obtained from a convenience cohort of children under 5 years of age presenting to a pediatric emergency department for evaluation of respiratory symptoms (demographics shown in Table 2). Children who tested positive for acute SARS-CoV-2 infection were excluded from this study. Children were categorized into 4 groups based on parental recall of COVID-19 mRNA vaccination and evidence of prior SARS-CoV-2 infection (presence of anti-nucleocapsid IgG in the VTM): No history of vaccination or evidence of SARS-CoV-2 infection (“Negative”), history of vaccination only (“Vaccine-only”), evidence for SARS-CoV-2 infection only (“Prior Infection”), and a history of both (“Vaccine/Infection”). We found that SARS-CoV-2-specific IgG levels were significantly higher in the Vaccine-only, Vaccine/Infection, and Prior Infection groups compared to the Negative group (Figure 2B), suggesting effective induction of SARS-CoV-2 specific IgG within the nasal mucosa by either vaccination or natural infection. Notably, levels of nasal IgG were significantly higher in children who were both vaccinated and had a prior SARS-CoV-2 infection compared to all other groups, indicating that COVID-19 mRNA vaccination likely boosts nasal IgG levels in participants previously infected with SARS-CoV-2.

**Figure 2.**
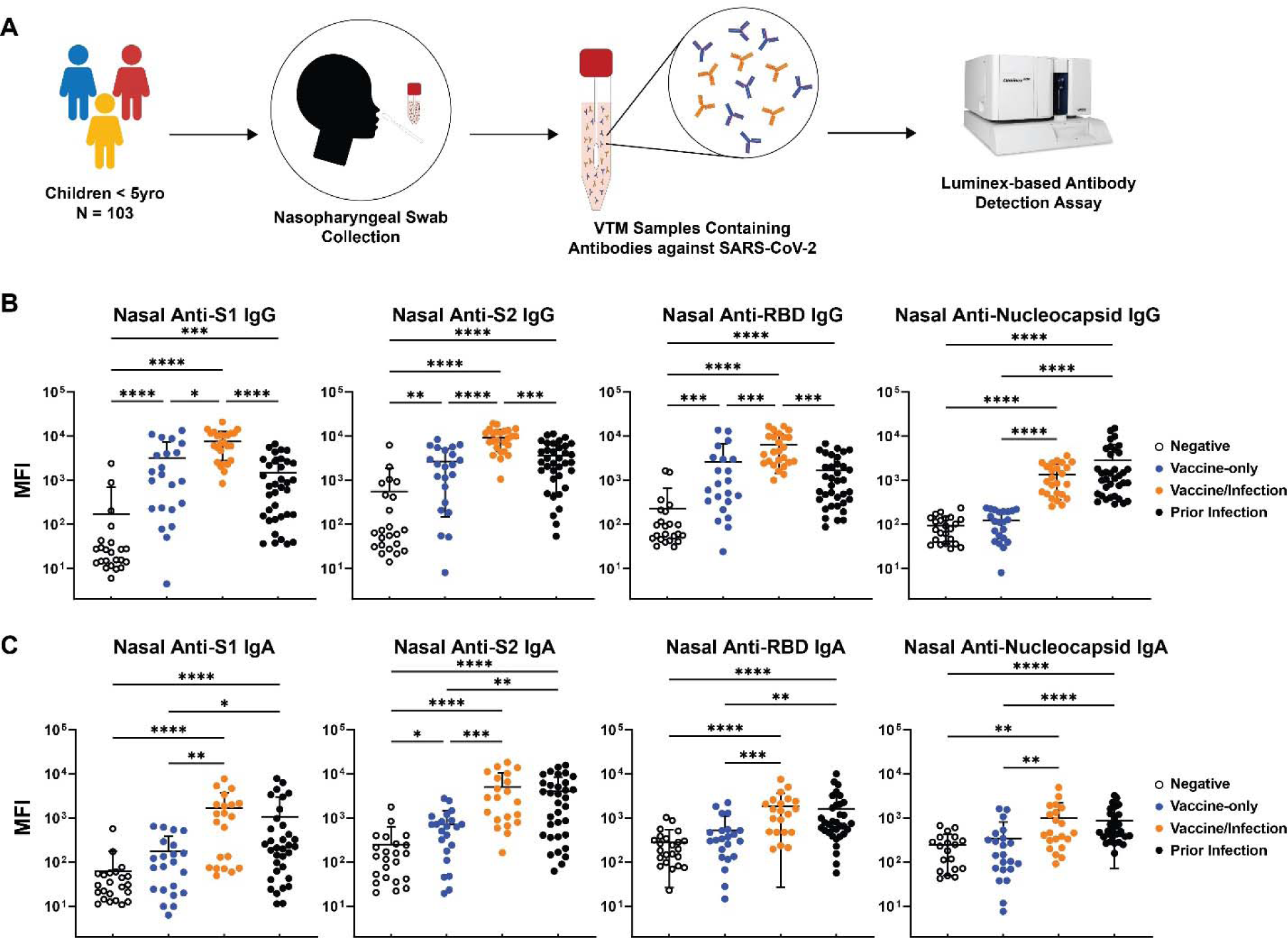
SARS-CoV-2 mRNA vaccination fails to induce anti-spike IgA in the nasal mucosa. **(A)** Schematic overview of study design and experimental procedures. **(B-C)** Nasal anti-S1, -S2, -RBD, and -Nucleocapsid IgG (B) and IgA (C) levels were plotted, and comparisons among four groups were conducted. Error bar represents the mean value and the standard deviation. Two-tailed Mann-Whitney U tests were performed, and statistical significance is defined as *p < 0.05, **p < 0.01, ***p < 0.001, and ****p < 0.0001.

**Table 2.** Characteristics of participants in emergency department convenience cohort.

|  | Negative<br>(N = 23) | Vaccine-only<br>(N = 22) | Vaccine/Infection<br>(N = 22) | Prior Infection<br>(N = 36) |
| --- | --- | --- | --- | --- |
| **Age (month)** |  |  |  |  |
| Minimum | 5 | 8 | 3 | 2 |
| Median (IQR) | 24 (24) | 18.5 (24) | 29 (25) | 25.5 (22) |
| Maximum | 47 | 57 | 53 | 58 |
| **Sex** |  |  |  |  |
| Female | 5 (21.7%) | 9 (40.9%) | 12 (54.5%) | 17 (47.2%) |
| Male | 18 (78.3%) | 13 (59.1%) | 10 (45.5%) | 19 (52.8%) |
| **Race** |  |  |  |  |
| White | 6 (26.1%) | 11 (50%) | 7 (31.8%) | 12 (33.3%) |
| Black/African American | 3 (13%) | 1 (4.5%) | 3 (13.6%) | 4 (11.1%) |
| Asian | 2 (8.7%) | 3 (13.6%) | 0 (0%) | 3 (8.3%) |
| Other/Unknown | 12 (52.2%) | 7 (31.8%) | 12 (54.5%) | 17 (47.2%) |
| **Ethnicity** |  |  |  |  |
| Hispanic | 5 (21.7%) | 0 (0%) | 6 (27.3%) | 10 (27.8%) |
| Not Hispanic | 12 (52.2%) | 13 (59.1%) | 8 (36.4%) | 16 (44.4%) |
| Unknown | 6 (26.1%) | 9 (40.9%) | 8 (36.4%) | 10 (27.8%) |
| **COVID-19 Vaccine Status** |  |  |  |  |
| Not vaccinated | 23 (100%) | 0 (0%) | 0 (0%) | 36 (100%) |
| Pfizer-BioNTech | 0 (0%) | 9 (40.9%) | 9 (40.9%) | 0 (0%) |
| Moderna | 0 (0%) | 9 (40.9%) | 8 (36.4%) | 0 (0%) |
| Not known | 0 (0%) | 4 (18.2%) | 5 (22.7%) | 0 (0%) |

In contrast, nasal anti-S1, anti-S2, and anti-RBD IgA levels were significantly higher in the Vaccine/Infection and Prior Infection groups than in both the Negative and Vaccine-only groups, and we were unable to detect a significant difference in anti-S1 or anti-RBD IgA levels between the Vaccine-only group and the Negative group, nor between the Vaccine/Infection group and the Prior infection group (Figure 2C). We did detect a small but significant increase in anti-S2 IgA levels between the Vaccine-only group and the Negative group, although we did not appreciate a significant increase in anti-S2 IgA between the Vaccine/Infection group and the Prior Infection group. Similar to results with saliva, these results indicate that despite the ability to induce mucosal IgG, the ability of COVID-19 mRNA vaccination to induce SARS-CoV-2 specific IgA in the nasal mucosa is quite limited.

### SARS-CoV-2 specific salivary and nasal antibodies trigger extensive spike-mediated neutrophil activation

Neutrophils are abundant in the nasal mucosa of healthy children, and exhibit a more activated phenotype than neutrophils in the adult nose following SARS-CoV-2 infection [11]. However, whether SARS-CoV-2 specific antibodies in the oronasopharynx have the ability to activate neutrophils following antigen exposure is not fully defined, and further, the role of mucosal IgA in this process remains to be determined. To examine whether mucosal antibodies induced by vaccination and/or natural infection have the ability to activate neutrophils and induce the formation of neutrophil extracellular traps (NET), we pooled saliva samples from healthy children with completed vaccine doses in the following groups: “Negative” (no prior infection or vaccination), “Vaccine-only” (vaccinated individuals without history of COVID-19) and “Vaccine/Infection” (vaccinated individuals with prior infection) (N=4 samples per pool) to obtain sufficient volumes of saliva to evaluate NET formation. We then mixed these pooled saliva samples with spike protein-coated beads to induce immune complex formation and added these mixtures to neutrophils isolated from four healthy individuals. We assessed neutrophil activation by quantification of the percentage of neutrophils that undergo NETosis (Figure 3A).

**Figure 3.**
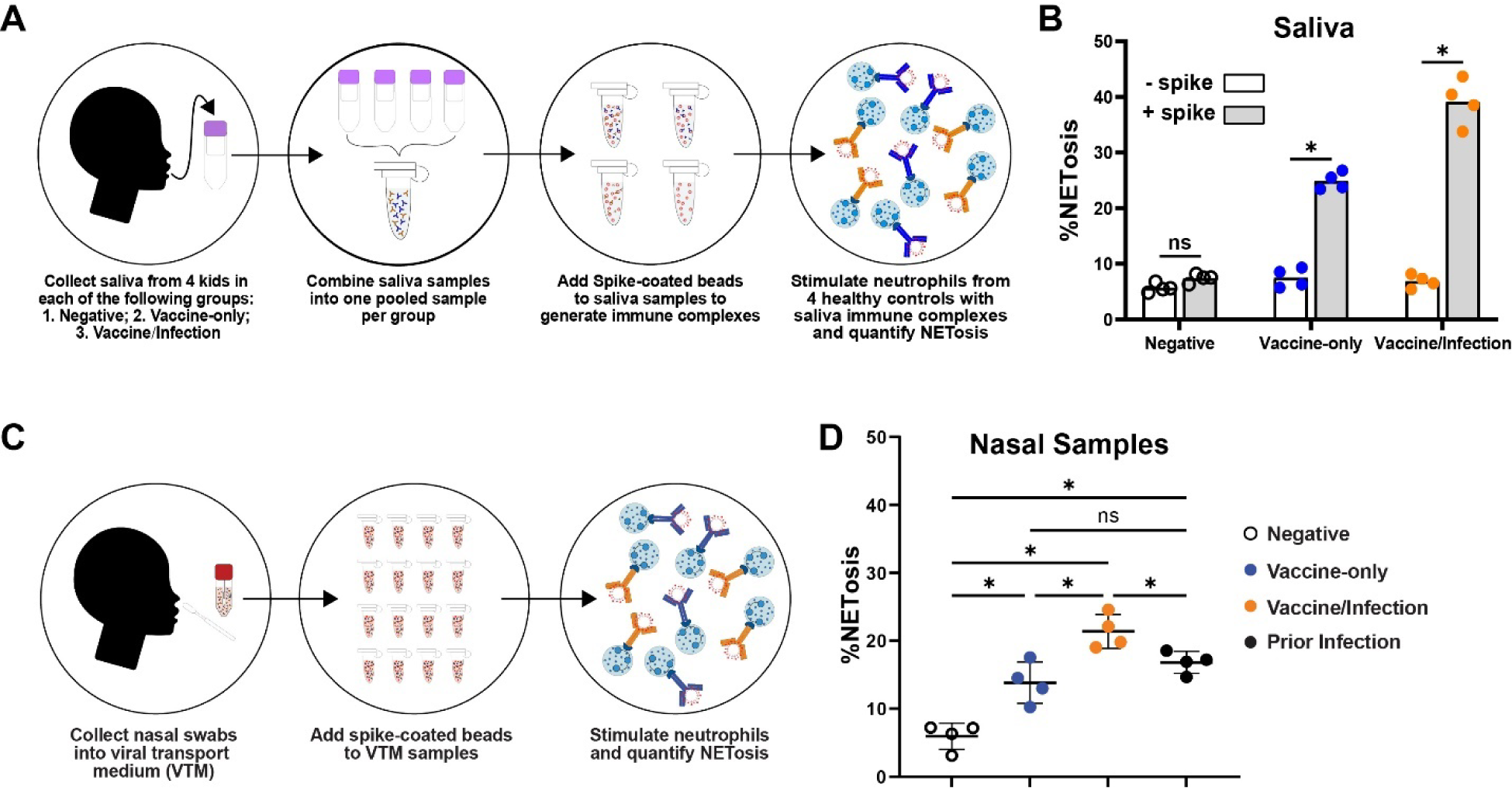
SARS-CoV-2 specific salivary and nasal antibodies trigger extensive spike-mediated neutrophil activation. **(A)** Schematic overview of spike-mediated NETosis assay using saliva pools. **(B)** Comparison of %NETosis in the presence (white bars) or absence (gray bars) of spike-coated beads in the Negative, Vaccine-only, and Vaccine/Infection pools. **(C)** Schematic overview of spike-specific NETosis assay using individual nasal samples (N=4 per group). **(D)** Percent NETosis of neutrophils stimulated by spike-coated beads with nasal samples from Negative, Vaccine-only, Vaccine/Infection, and Prior Infection groups. Error bar represents the mean value and the standard deviation. Two-tailed Mann-Whitney U tests were performed, and statistical significance is defined as *p < 0.05.

None of the sample pools induced NETosis in the absence of spike protein, but we observed significant increases in NETosis following the addition of spike protein to the Vaccine-only pool and the Vaccine/Infection pool, but not from the Negative pool (Figure 3B), consistent with the presence of antibodies with the ability to induce anti-spike immune complexes in these pools. Interestingly, the level of NETosis was higher in the Vaccine/Infection pool than in the Vaccine-only pool, likely reflecting the higher levels of salivary anti-SARS-CoV-2 antibodies, although the analysis of a single pool limits our ability to evaluate significance across pools assembled from the different groups. To address this potential limitation, we compared the ability of a subset of nasal samples (N=4 per group) to induce NETosis following exposure to spike protein-coated beads (Figure 3C). Antibody levels for each individual sample used in this assay are shown in Figure S3. The induction of NETosis was significantly higher in Vaccine-only, Vaccine/Infection, and Prior Infection groups than in the Negative group, and we observed significantly higher levels of NETosis in the Vaccine/Infection group than in all other groups (Figure 3D). Taken together, these results confirm that higher levels of antibodies observed within the oronasal mucosa of vaccinated children with a prior SARS-CoV-2 infection are associated with an enhanced neutrophil activation, likely signifying functional importance.

### Spike-specific IgA in saliva acts as a key inducer of neutrophil activation

To better understand which subclass of antibodies drives the NETosis observed in salivary samples, we depleted either IgG, IgA, or both from pooled saliva samples and evaluated neutrophil activation following the addition of spike protein-coated beads (Figure 4A). We found that depletion of either IgG or IgA from the Vaccine-only or Vaccine/Infection pools significantly inhibited NETosis and that NETosis was eliminated by depletion of both IgG and IgA (Figure 4B and 4C). Thus, both IgG and IgA contribute to the ability of oronasal mucosal antibodies to induce SARS-CoV-2 specific neutrophil activation in children exposed and/or immunized to the virus. One inconsistency we noted is that IgA depletion in Vaccine-only saliva pool significantly inhibited the neutrophil NET formation, even though we did not observe significant induction of mucosal IgA by vaccination alone. We believe that a non-specific ability of IgA in saliva to induce NET formation is unlikely, given that saliva from children without a history of COVID-19 did not induce NET formation prior to vaccination. Rather, we suspect that vaccination alone does result in low levels of mucosal IgA potentially from passive transport from serum and that we were unable to detect significant differences in levels of nasal IgA between the unvaccinated and vaccinated groups given our sample size limitations. Future studies with larger sample sizes will be necessary to definitively answer this question.

**Figure 4.**
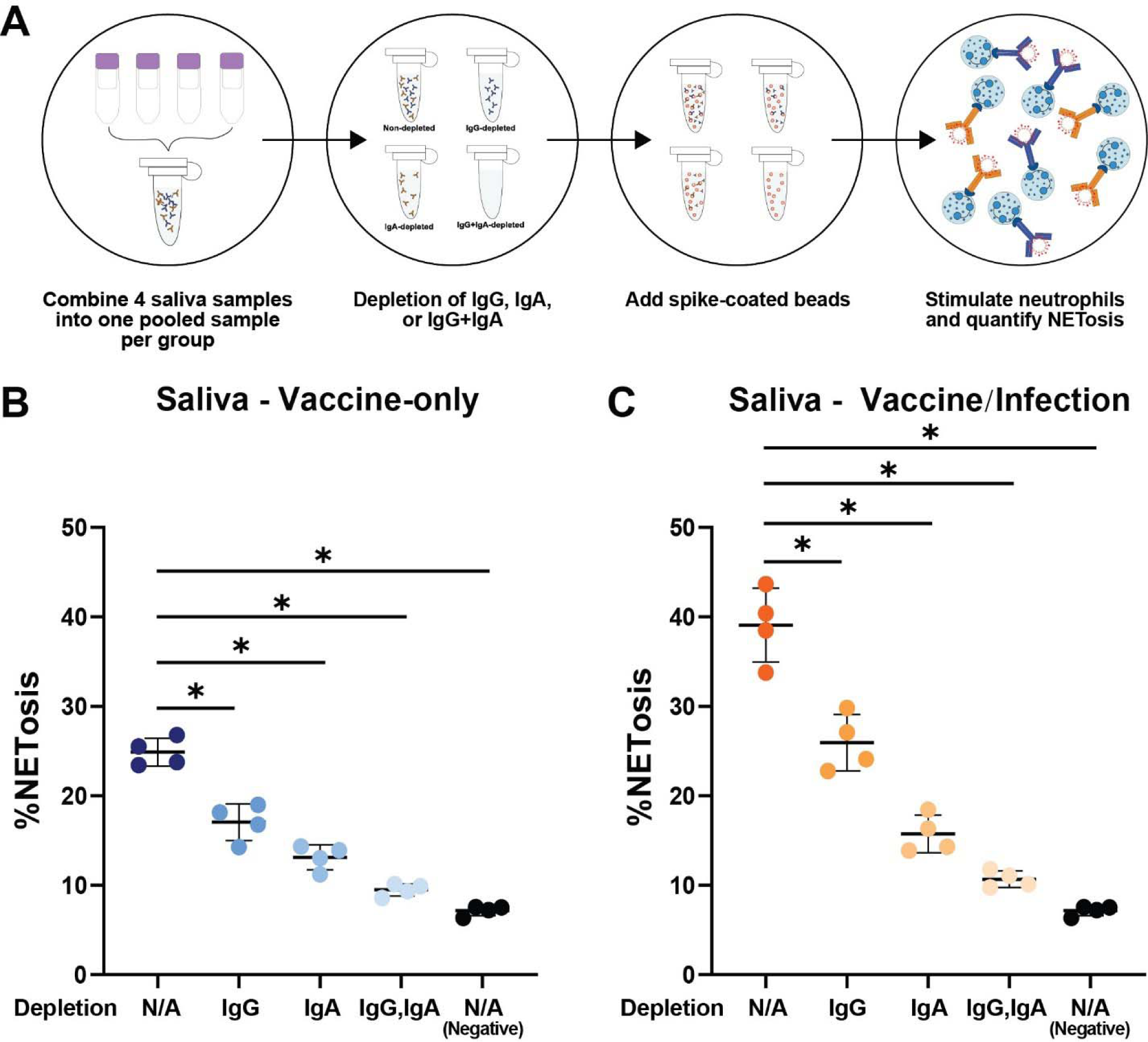
Depletion of mucosal antibodies interferes with the neutrophil activation induced by saliva pools from individuals in the Vaccine-only and Vaccine/Infection group. **(A)** Schematic overview of antibody depletion assay in salivary samples. **(B-C)** End-point percentage of NETs released from neutrophils stimulated with saliva from the Vaccine-only pool (B) and the Vaccine/Infection pool (C) following depletion of IgG, IgA, or both IgG and IgA. Black dots represent NETs released from neutrophils stimulated with the Negative saliva pool in the presence of spike-coated beads. Error bar represents the mean value and the standard deviation. Two-tailed Mann-Whitney U tests were performed, and statistical significance is defined as *p < 0.05.

## DISCUSSION

In June 2022, the FDA granted approval for the administration of the COVID-19 mRNA vaccine to children aged 6 months to 5 years, however over 95% of children have been exposed to SARS-CoV-2 based on national serological surveillance testing [12]. Thus, it is essential to conduct a thorough evaluation of both systemic and mucosal humoral responses triggered by immunization in children under 5 years of age with and without prior infection.

The robust anti-spike IgG and IgA responses we observed in serum post-vaccination aligned with the broader consensus regarding the efficacy of current mRNA vaccine in inducing systemic immunity. In addition, we revealed that vaccination induced mucosal IgG responses in children, though hybrid immunity induced the highest level of SARS-CoV-2 specific IgG. Our observation of close correlation between systemic and mucosal IgG levels is consistent with models in which IgG accumulates in the mucosa as the result of passive transport from the circulatory system [13]. In contrast, our study highlighted the limited ability of these vaccines to generate mucosal IgA responses, and confirms that mucosal IgA production in the oronasopharynx can be largely independent of systemic IgA responses [14]. IgA is recognized as an important factor in mucosal immunity regarding its role in neutralizing pathogens, particularly in the gastrointestinal tract and the upper airways [15]. Notably, mucosal IgA has been identified as a critical antibody type protecting against SARS-CoV-2 infection [16,17] and correlates with reduced viral infectivity *in vitro* [18]. Our findings raised questions about the completeness of protection conferred by the current immunization strategies, although the exact function of viral-specific mucosal IgA still requires further investigation.

Another crucial aspect of our study involves the exploration of mucosal antibody-induced neutrophil activation, as demonstrated by the assessment of neutrophil extracellular traps induced by salivary and nasal samples from infected and/or immunized individuals. Neutrophils have been shown to release NETs as an antimicrobial defense at the mucosa, helping to clear pathogens to prevent more severe infection and disease [19,20]. Also, children have abundant neutrophils in their airways, which may contribute to the rapid viral clearance and mild disease observed in children [11,21,22]. In our study, we found that vaccine and infection-induced mucosal antibodies were likely generating immune complexes upon spike protein challenge, resulting in enhanced NET formation. Furthermore, we identified the critical role of mucosal IgA in driving spike-mediated NETosis, suggesting the generation of SARS-CoV-2 specific mucosal IgA is a key component for providing enhanced protection against subsequent infections. While mucosal IgA immune complexes are the most potent inducer of NETs, IgG immune complexes were also able to induce NETs, albeit to a lesser degree, supporting that vaccination, through induction of mucosal IgG, provides some degree of mucosal protection which may contribute to more rapid clearance of virus in vaccinated as compared to unvaccinated individuals [23]. Vaccination in previously infected individuals provided the most abundant SARS-CoV-2 specific IgG, emphasizing the potential importance of continued vaccination efforts in this population.

Our study was limited by a relatively small sample size, however, we overcame numerous challenges in obtaining blood and mucosal samples from young pediatric cohorts with low rates of vaccination. Thus, this sized cohort is substantial enough to offer meaningful insights. Further, we utilized non-invasive samples such as saliva and nasopharyngeal swabs to demonstrate how mucosal immunity compared with systemic immunity.

In conclusion, our study confirms the ability of COVID-19 mRNA vaccines to induce mucosal in addition to systemic IgG in previously uninfected young children. However, the limited generation of mucosal IgA responses following vaccination underscores a potential area for improvement in current vaccination strategies for this specific demographic. Further research is warranted to explore alternative vaccine formulations or strategies that may enhance mucosal immunity in young children, contributing to more comprehensive protection against SARS-CoV-2.

## Data Availability

All data produced in the present study are available upon reasonable request to the authors.

## ACKNOWLEDGEMENT

Study design/funding: B.H.H., L.M.Y.; Data acquisition and analysis: Y.T., B.P.B., Z.N.S., E.B., S.C.; Patient consent and sample collection: W.I.G., M.D., M.A.F.L., A.F., M.D., R.L.W., J.G., J.S., Y.T.; Manuscript writing: Y.T.; Figure generation: Y.T., B.P.B.; Supervision: D.R.W., B.H.H., L.M.Y.; All authors reviewed and approved the final version of the manuscript.

We express extreme gratitude to all of the young children and families who participated in our study.

---

**Figure S1.**
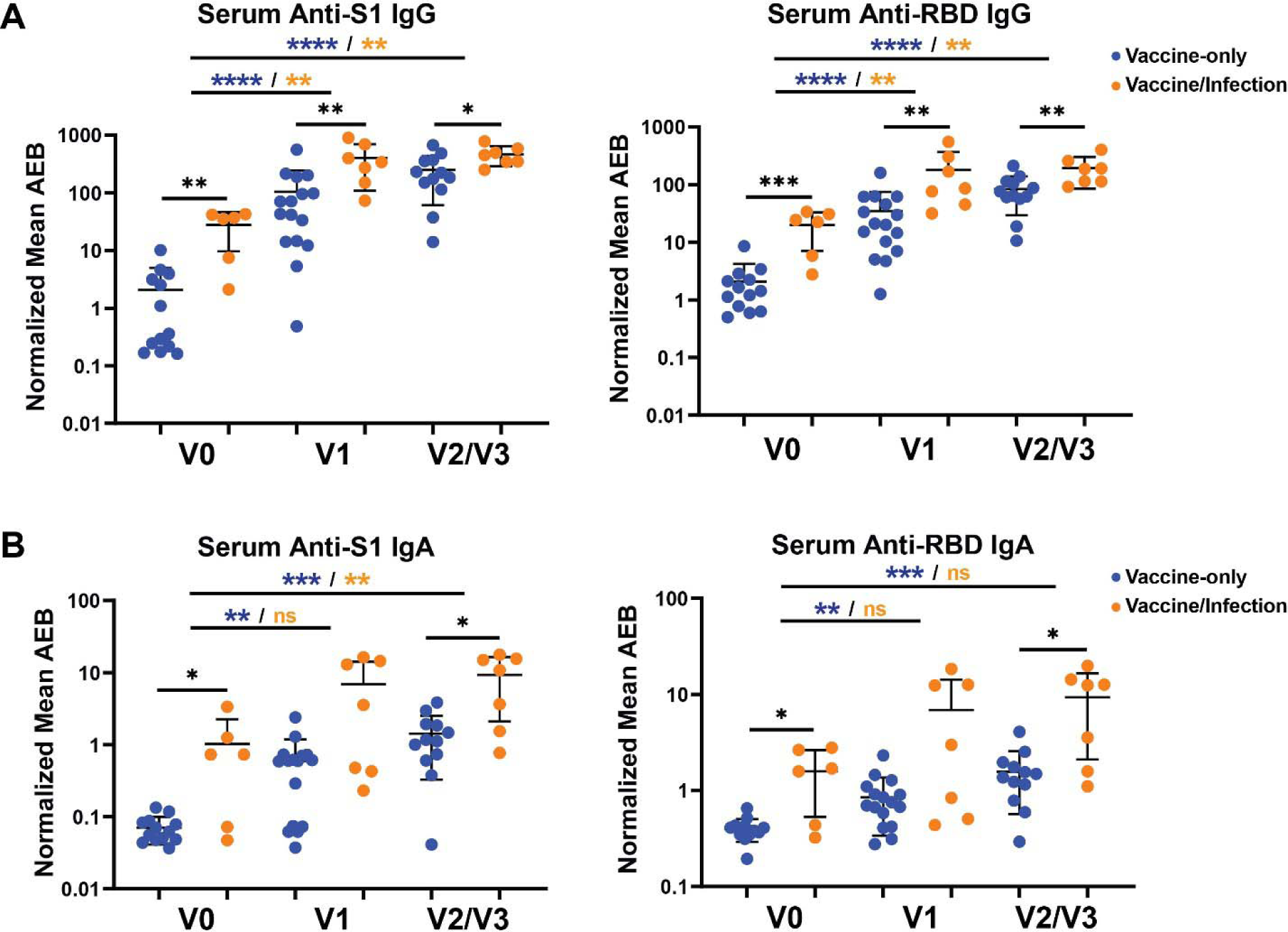
Vaccination induces anti-S1/-RBD IgG and IgA in the serum. **(A)** Serum anti-S1 (left) and anti-RBD (right) IgG levels are shown. **(B)** Serum anti-S1 (left) and anti-RBD (right) IgA levels are shown. Differences between groups are shown as black asterisks. Differences between time points within groups are shown as blue or orange asterisks. Error bar represents the mean value and the standard deviation. Two-tailed Mann-Whitney U tests were performed, and statistical significance is defined as *p < 0.05, **p < 0.01, ***p < 0.001, and ****p < 0.0001.

**Figure S2.**
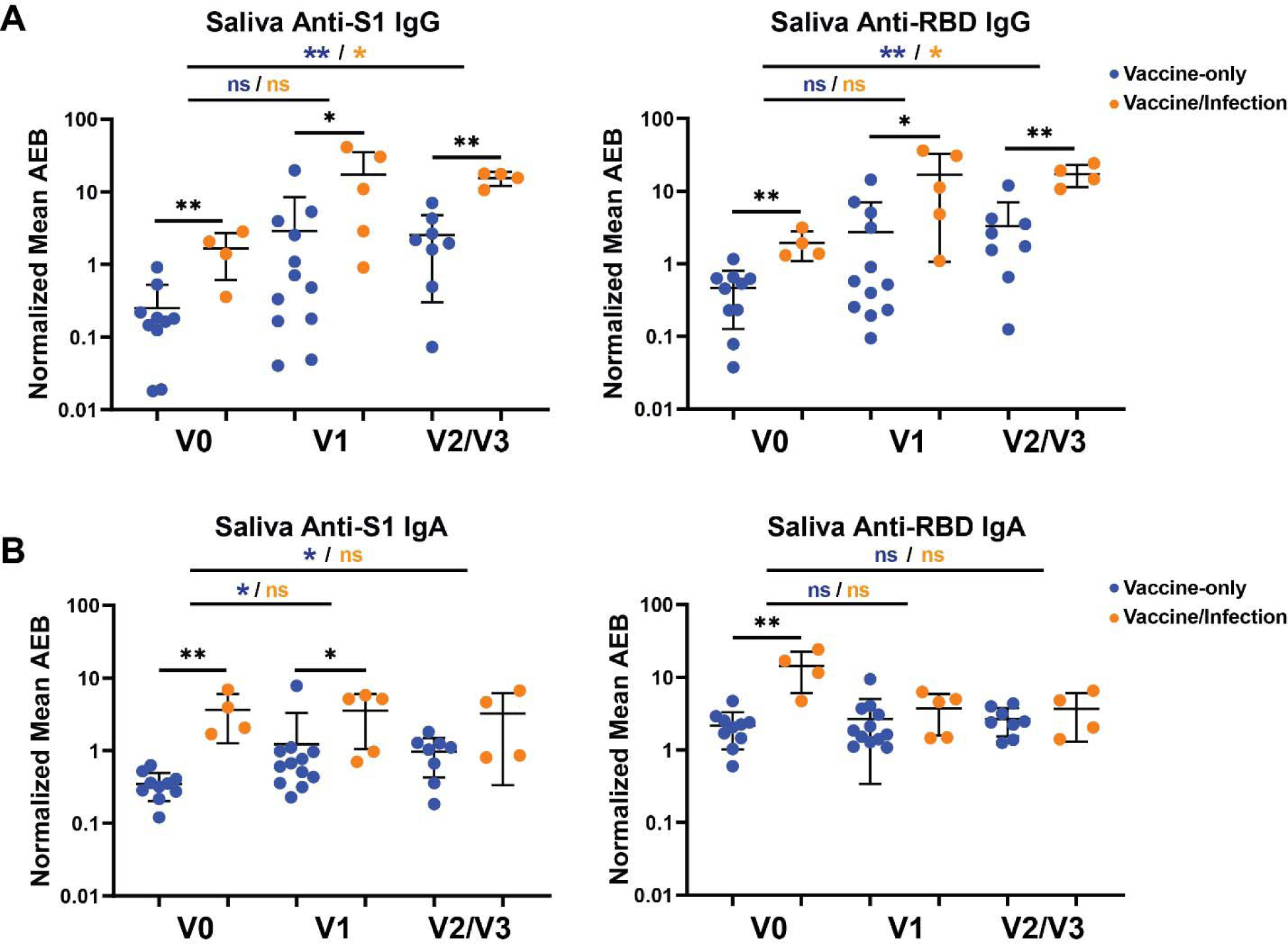
Vaccination induces salivary anti-S1/-RBD IgG but not IgA. **(A)** Saliva anti-S1 (left) and anti-RBD (right) IgG levels are shown. **(B)** Saliva anti-S1 (left) and anti-RBD (right) IgA levels are shown. Differences between groups are shown as black asterisks. Differences between time points within groups are shown as blue or orange asterisks. Error bar represents the mean value and the standard deviation. Two-tailed Mann-Whitney U tests were performed, and statistical significance is defined as *p < 0.05 and **p < 0.01.

**Figure S3.**
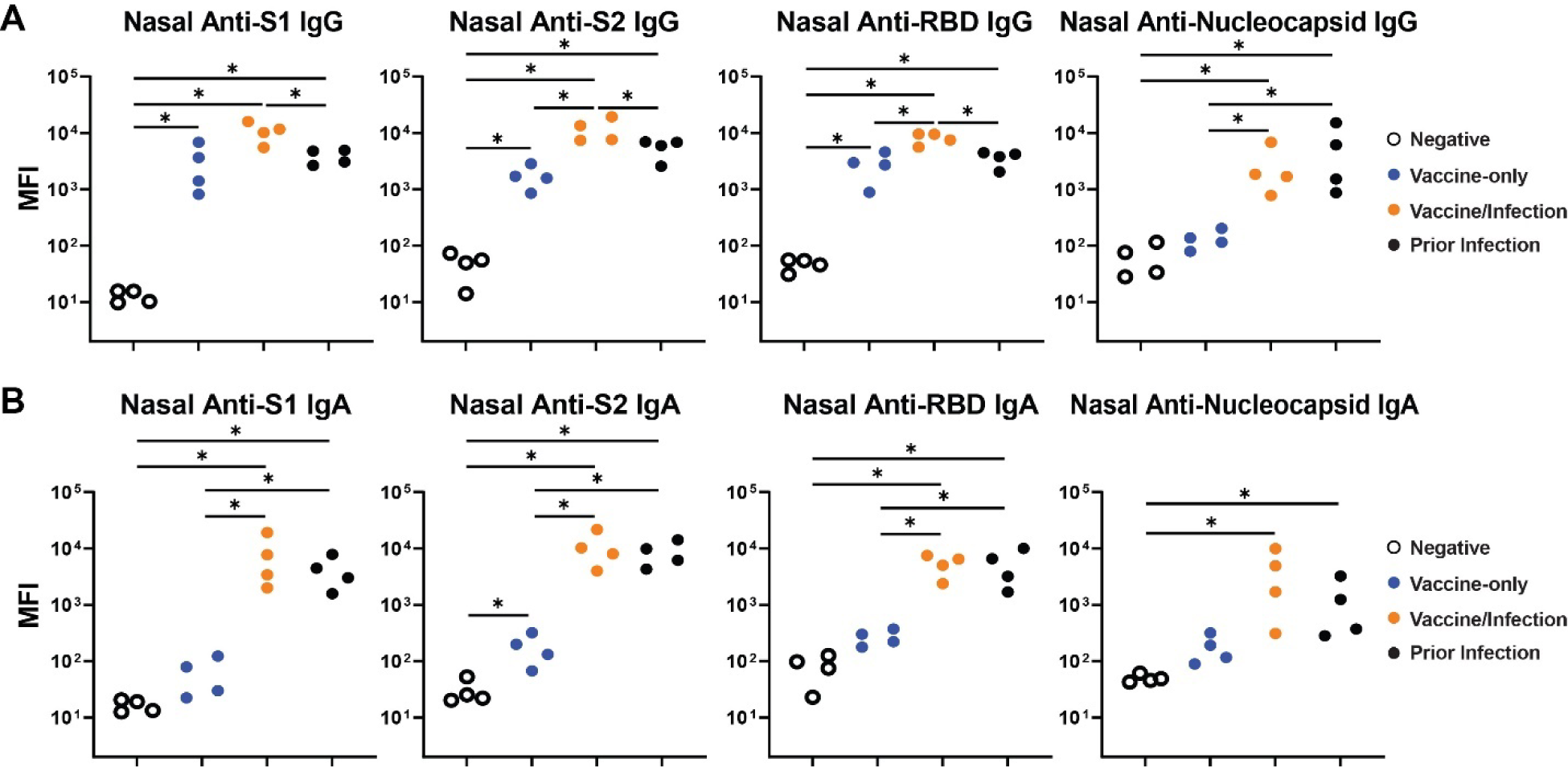
Levels of SARS-CoV-2 specific IgG and IgA in nasal samples evaluated in Figure 3D. Levels of Anti-S1, Anti-S2, Anti-RBD, and Anti-Nucleocapsid IgG **(A)** and IgA **(B)** in the 4 selected nasal samples from each group used for NETosis assay. Two-tailed Mann-Whitney U tests were performed, and statistical significance is defined as *p < 0.05.

